# Projected prevalence and number of persons with diagnosed type 2 diabetes by socioeconomic position in Germany between 2022 and 2060

**DOI:** 10.64898/2026.11.12.26367900

**Authors:** Katharina Piedboeuf-Potyka, Ramona Hering, Mandy Schulz, Malwina Mackowiak, Ralph Brinks, Annika Hoyer, Marielle Wirth, Thaddäus Tönnies

**Affiliations:** Institute for Biometrics and Epidemiology, German Diabetes Center (DDZ), Leibniz Center for Diabetes Research at the Heinrich Heine University, 40225 Düsseldorf, Germany; Department of Healthcare Analyses, Central Research Institute for Ambulatory Care in Germany, 10587 Berlin, Germany; Department of Mathematics, Informatics, Technology, Koblenz University of Applied Sciences, RheinAhrCampus, 53424 Remagen, Germany; Chair for Medical Biometry and Epidemiology (MBE), Witten/Herdecke University, Faculty of Health/School of Medicine, 58448 Witten, Germany; Biostatistics and Medical Biometry, Medical School OWL, Bielefeld University, 33615 Bielefeld, Germany; Department of General Pediatrics, Neonatology and Pediatric Cardiology, Medical Faculty and University Hospital Düsseldorf, Heinrich Heine University Düsseldorf, 40225 Düsseldorf, Germany

**Keywords:** Type 2 Diabetes, Social Inequalities, Germany, Health Inequalities, Prevalence

## Abstract

**Background:** Current projections of the prevalence and number of persons with type 2 diabetes by socioeconomic position (SEP) are lacking for Germany. This study aims to project age-specific type 2 diabetes prevalence in Germany by sex and SEP between 2022 and 2060.

**Methods:** We used a partial differential equation (PDE) based on the illness-death model to project age-specific type 2 diabetes prevalence by sex and SEP between 2022 and 2060. The PDE describes the relationship between prevalence, incidence, all-cause mortality and the mortality rate ratio (MRR) of individuals with vs. without type2 diabetes considering age, calendar year and SEP. SEP was measured by quintiles of the German Index of Socioeconomic Deprivation (GISD). Baseline prevalence, incidence, and MRR in 2022 were derived from statutory health insurance data (∼85% of the German population). Projected prevalence was multiplied by population projections from the German Federal Statistical Office to estimate the number of persons with diabetes.

**Results:** Type 2 diabetes prevalence increases with age, peaking between 80 and 90 years. Between 2022 and 2060, age-specific prevalence is projected to increase for both sexes across all SEP groups. SEP-related disparities persisted with largest differences between the highest (5th GISD quintile) and lowest (1st GISD quintile) SEP group. The number of persons with type 2 diabetes is projected to rise from 8.4 million in 2022 to between 11.2 and 19.2 million in 2060.

**Conclusions:** Between 2022 and 2060, age-specific type 2 diabetes prevalence is projected to increase with increasing socioeconomic inequalities.

## INTRODUCTION

The global number of persons with diabetes has increased over the past decade,^1^ a trend that is also reflected in Germany, where type 2 diabetes poses a growing public health challenge.^2, 3^

As in many other countries, the majority of diabetes cases in Germany are classified as type 2, and the number of affected individuals has been rising steadily over recent years.^3^ Previous projections suggest an increase in type 2 diabetes prevalence from 6.8 million persons in 2015 to between 10.9 and 14.2 million persons in 2040.^2^ In line with these projections, the prevalence of type 2 diabetes in Germany was estimated at 9.5% in 2023.^3^

Several studies indicate that the projected type 2 diabetes prevalence largely depends on temporal trends in the incidence of type 2 diabetes.^4^ While extrapolating current incidence trends provides a foundation for estimating future prevalence, it is also essential to consider that these future prevalence trends are not socioeconomically neutral. The burden of type 2 diabetes increasingly concentrates in populations with lower socioeconomic position (SEP). Higher prevalence is consistently observed in socially deprived areas, where material disadvantage accumulates and reinforces health inequities.^5^ Social deprivation refers to the relative lack of socioeconomic and material resources and the associated social and spatial burdens within an area.^6^ For instance Evans et al.^7^ have discovered that the prevalence of type 2 diabetes varied by deprivation, and that persons in the highest deprivation category were 1.6 (95% confidence interval 1.4–1.8) times more often affected by type 2 diabetes compared to persons in the lowest deprivation category.^7^ In addition, a systematic review indicated that the SEP seems to be a risk factor for diabetes complications.^8^ Although there is a strong association between SEP and the risk of type 2 diabetes and numerous studies project the future prevalence of type 2 diabetes in Germany,^2, 4, 9^ none of these projections take the SEP into account. Contrary to the evidence that future prevalence depends on temporal trends in the type 2 diabetes incidence, previous projections did not base their projections on observed trends in the incidence.^4, 9^ Moreover, the data that informed previous projections rely on input data dating back to 2010.^4, 9^ For these reasons we aimed to project the age- and sex-specific type 2 diabetes prevalence by SEP in Germany between 2022 and 2060 based on data available from 2022, including current temporal trends in type 2 diabetes incidence.

## METHODS

### Projection model

As in previous studies,^2, 4^ we applied an illness-death model to project the future age- and sex-specific type 2 diabetes prevalence between 2022 and 2060. The model describes the relationship between the prevalence *p*, the incidence *i*, the all-cause mortality *m*, and the mortality rate ratio (MRR) for individuals with vs. without type 2 diabetes. Each of these quantities depend on the calendar year and age. The temporal change in prevalence, ∂*p*, is described by the following partial differential equation (PDE)

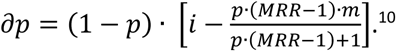

The projection of type 2 diabetes prevalence is performed by solving the PDE through numerical integration. Further information on the illness-death-model can be found in previous publications.^2, 4, 10^

### Data sources and input data

To solve the PDE, different input data are required: age-specific prevalence and incidence of type 2 diabetes in the first year of the projection period, all-cause mortality, and MRR. We used publicly available data from previous studies and federal statistical offices on age- and sex-specific type 2 diabetes prevalence, incidence and mortality, which are representative for Germany.^11^ To project future trends in type 2 diabetes prevalence across different socioeconomic groups, we extended previous applications of the prediction model by incorporating SEP-specific data at the county level. This was also accomplished by incorporating SEP-specific input values of all-cause mortality, prevalence and incidence from previous studies^11^ and federal statistical offices.^12^ Consequently, the temporal change in prevalence depends not only on calendar year, age and sex, but also on SEP. Initial values for age- and sex-specific type 2 diabetes prevalence and incidence by SEP were obtained from previous studies using data from German statutory health insurance covering approximately 85% of the German population. The data, which were aggregated at the county level, included all statutory health insurance providers and age groups.^11, 13^

Since the original data sources^3^ did not contain information on SEP, previous studies^11, 13^ linked these data to the publicly available area-based German Index of Socioeconomic Deprivation (GISD). The GISD is based on aggregated regional data to quantify SEP at the county level, ranges from 0 (highest SEP) to 1 (lowest SEP), and combines information on education, occupation, and income.^6^ The distribution of the GISD between 2014 and 2019 is shown in Supplement 1.

We used the age-, sex- and GISD-specific prevalence for 2022 from Wirth et al.^13^ as the starting prevalence for the projection. For type 2 diabetes incidence in the starting year 2022, we combined data on age- and sex-specific incidence in the general population from 2022, provided by the Central Research Institute for Ambulatory Health Care in Germany (Zi)^3^, with sex-specific and age-standardized incidence ratios (SIR) from Piedboeuf-Potyka et al.^11^ to obtain age- and sex-specific type 2 diabetes incidence by quintiles of the GISD. More specifically, the SIR is the age-standardized ratio of GISD-specific incidence and the incidence of the general population. Hence, multiplying the age- and sex-specific incidence of the general population by the sex-specific SIR results in age- and sex-specific incidence estimates for the different GISD groups. Since SIR estimates were only available until 2019, we updated the previous analysis with data until 2022 using the same procedures described in Piedboeuf-Potyka et al.^11^ Results of the updated analysis are reported in Supplement 2. All-cause current and future mortality by sex and GISD were based on data from official population projections from the German Federal Statistical Office.^12, 14^ Estimates for input values of the age- and sex-specific MRRs were taken from Schmidt et al.^15^

### Projection scenarios

We projected five scenarios of age-, sex- and GISD-specific type 2 diabetes prevalence between 2022 and 2060. For all projections, we assumed a constant GISD quintile distribution based on the most recent data from 2019. We further assumed annual decreases in the MRR of 0.60% for women and 0.72% for men across all scenarios, drawing on evidence from Denmark.^16^

Because GISD-specific MRR estimates were unavailable, we applied the same age- and sex-specific MRRs as in Schmidt et al.^15^ for all GISD groups.

The scenarios differed regarding future trends in age-, sex-, and GISD-specific type 2 diabetes incidence. In scenario 1, we assumed constant incidence between 2022 and 2060. Scenarios 2-3 represent increasing, and scenarios 4-5 decreasing incidence trends until 2027 (5-year in- or decrease) and 2032 (10-year in- or decrease), respectively (Supplement 2). Specifically, scenarios 2 and 3 extrapolated non-linear trends observed between 2014 and 2022, thereby capturing the observed increase in more recent years. For scenarios 4 and 5, we used the linear decrease observed in the pre-pandemic period 2014-2019. Details and visualizations of the scenarios are shown in Supplements 3–5.

To calculate the number of persons with type 2 diabetes, we multiplied the projected age- and sex-specific prevalence with the age- and sex-specific population projections from the German Federal Statistical Office.^12^ For the population projection, we assumed moderate developments in future life expectancy at birth, birth rates, and migration. We only considered the population aged 18 years and older, given the negligible type 2 diabetes prevalence in younger age groups.^4^

In the main analyses, we assume that migrants have the same prevalence as the resident population and therefore do not account for potential differences in prevalence among immigrants and emigrants. To assess the robustness of our results to potentially different prevalence levels among immigrants and emigrants, we considered two alternative scenarios in sensitivity analyses. In the first scenario, we assumed a threefold higher prevalence among persons immigrating to Germany and a prevalence one-third that of the resident population among persons emigrating from Germany. In the second scenario, these assumptions were reversed, with a prevalence one-third that of the resident population among immigrants and a threefold higher prevalence among emigrants. These assumptions are hypothetical, as empirically observed prevalence estimates for these groups are not available. The projected numbers of immigrants and emigrants were based on the population projections of the Federal Statistical Office.^12^

## RESULTS

### Projected prevalence

Figure 1 shows the age-specific prevalence of type 2 diabetes in Germany by sex and quintiles of the GISD for the five scenarios in the years 2022, 2040 and 2060. All scenarios project an increasing age-specific prevalence, particularly among older age groups, followed by a decline at the oldest ages.

**Figure 1:**
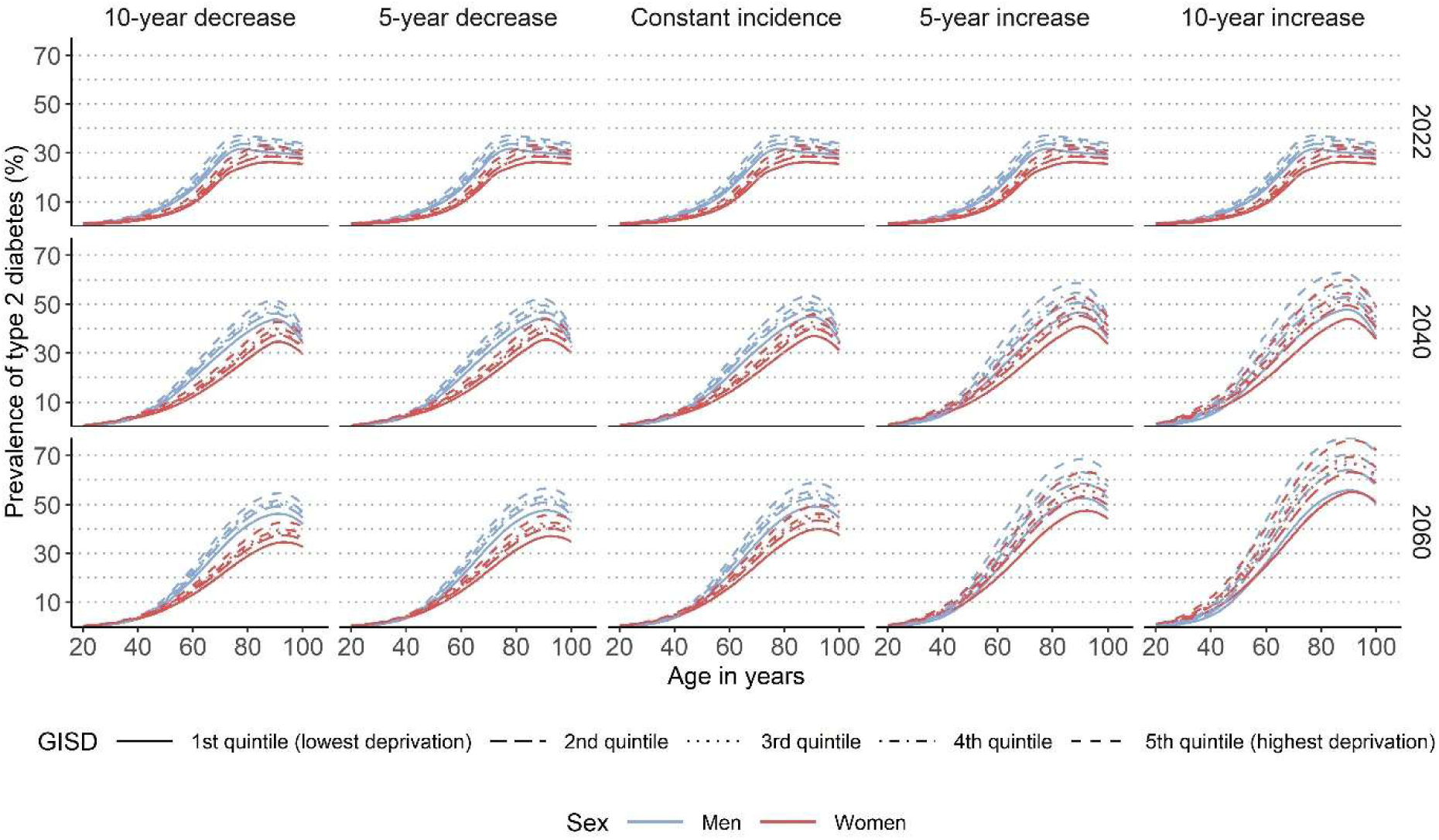
Projected age-specific prevalence of type 2 diabetes in Germany in 2022, 2040 and 2060 among men (blue) and women (red) by quintiles of the German Index of Socioeconomic Deprivation (GISD), shown for all five scenarios. Decreasing and increasing scenarios incorporate declining or rising type 2 diabetes incidence until 2027 for the 5-year de- or increase and 2032 for the 10-year de- or increase, respectively, while the constant scenario assumes stable incidence from 2022 to 2060. Scenarios with increasing type 2 diabetes incidence extrapolate the non-linear trends observed between 2014 and 2022, capturing the recent increase, whereas decreasing scenarios extrapolate the long-term linear decrease in incidence.

Moreover, the highest prevalence is consistently observed in these older populations. In the scenario with a 5-year increase in incidence the prevalence peaks at ∼83 years, lying between 26–37% among all GISD groups and both sexes in 2022, while in 2060 the prevalence peaks at ∼91 years, lying between 47–68% among GISD groups and both sexes.

Higher socioeconomic deprivation was associated with higher age-specific type 2 diabetes prevalence with the largest difference between the highest and lowest GISD group. In the scenario with a 5-year increase in type 2 diabetes incidence, the projected age-specific prevalence in 2060 is 68% for men and 63% for women in the highest GISD group, and 52% for men and 47% for women in the lowest GISD group, corresponding to the peak prevalence at about age 90. It is also observed that the prevalence of men is higher than that of women in early years, but that these differences diminish in the increasing incidence scenarios, reflecting the observed steeper increase in women in recent years (Supplement 5).

### Projected number of persons with type 2 diabetes

Figure 2 shows the projected number of persons with type 2 diabetes in Germany between 2022 and 2060 for all five scenarios by sex, summed across all GISD groups. In all scenarios, the sex-specific number of persons with type 2 diabetes increases between 2022 and 2060. Notably, in scenarios with increasing incidence, the projected number of women with type 2 diabetes surpasses that of men.

**Figure 2:**
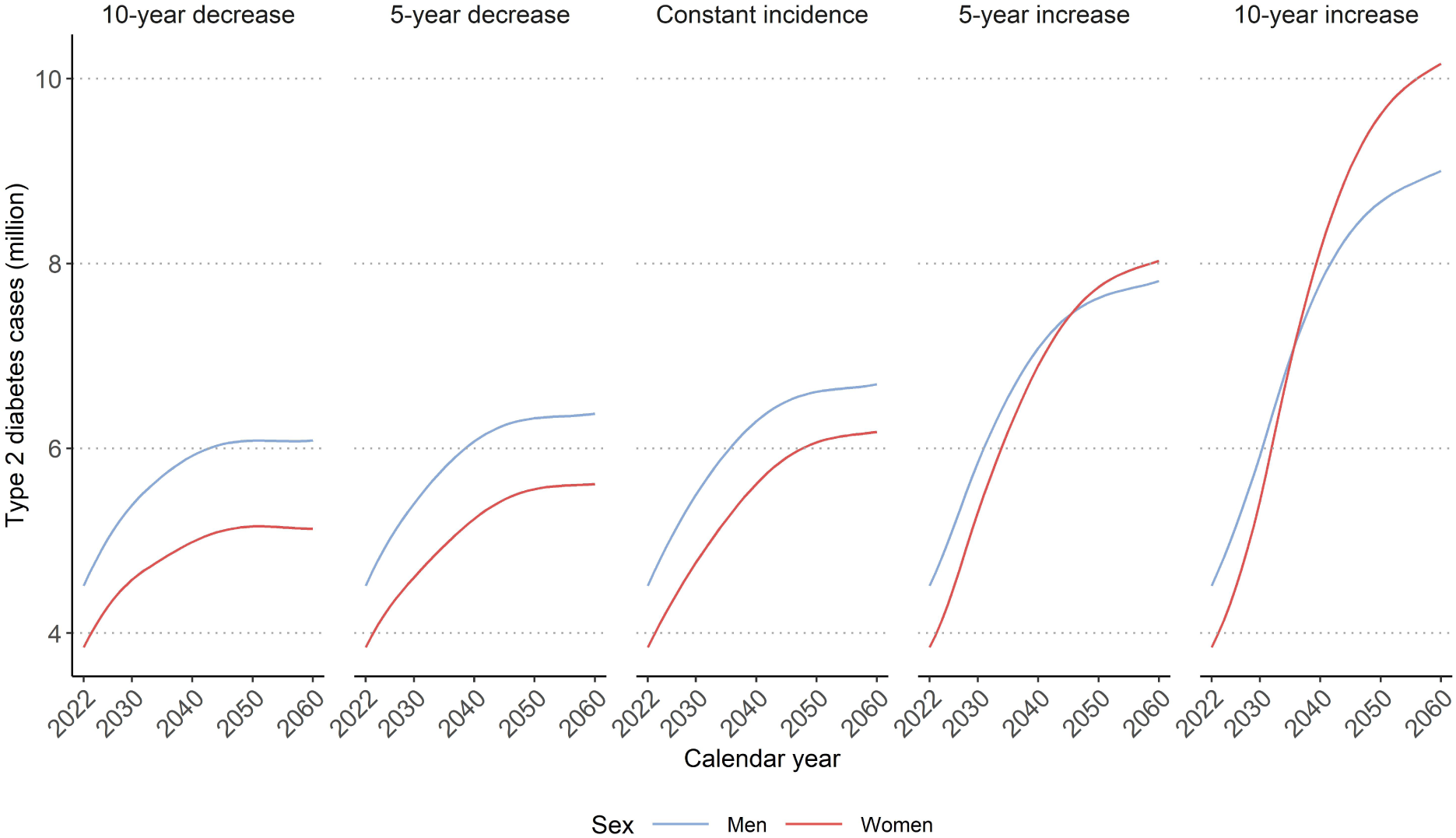
Projected number of type 2 diabetes cases in Germany between 2022 and 2060 summed across all quintiles of the German Index of Socioeconomic Deprivation (GISD), by sex shown for all five scenarios. Decreasing and increasing scenarios show declining or rising type 2 diabetes incidence until 2027 (5-year de- or increase) and 2032 (10-year de- or increase), respectively, while the constant scenario assumes stable incidence from 2022 to 2060. Scenarios with increasing type 2 diabetes incidence extrapolate the non-linear trends observed between 2014 and 2022, capturing the recent increase, whereas decreasing scenarios extrapolate the long-term linear decrease in incidence.

Table 1 shows the total number of type 2 diabetes cases by sex for 2022, 2040, and 2060. In the scenario with a 5-year increase in incidence, the total number of cases among women increased from approximately 3.8 million in 2022 to 8.0 million in 2060. A comparable increase was observed among men, with numbers rising from about 4.5 million to 7.8 million over the same period, resulting in a total increase from 8.3 million cases in 2022 to 15.8 million in 2060.

**Table 1:** Prevalence and total number of type 2 diabetes cases in Germany in 2022, 2040 and 2060 by sex, shown for all five scenarios. Decreasing and increasing scenarios show declining or rising type 2 diabetes incidence until 2027 and 2032, respectively, while the constant scenario assumes stable incidence from 2022 to 2060. Scenarios with increasing type 2 diabetes incidence extrapolate the non-linear trends observed between 2014 and 2022, capturing the recent increase, whereas decreasing scenarios extrapolate the long-term linear decrease in incidence.

| Sex | Calendar year | 10-year decrease | 5-year decrease | Constant incidence | 5-year increase | 10-year increase |
|---|---|---|---|---|---|---|
|  |  | **Prevalence (%)** |  |  |  |  |
| Women | 2022 | 9.0 | 9.0 | 9.0 | 9.0 | 9.0 |
| Men | 2022 | 10.9 | 10.9 | 10.9 | 10.9 | 10.9 |
| **Total** | 2022 | 9.9 | 9.9 | 9.9 | 9.9 | 9.9 |
| Women | 2040 | 11.6 | 12.1 | 13.0 | 16.0 | 18.9 |
| Men | 2040 | 14.2 | 14.6 | 15.1 | 17.0 | 18.7 |
| **Total** | 2040 | 12.9 | 13.3 | 14.0 | 16.5 | 18.8 |
| Women | 2060 | 12.2 | 13.3 | 14.7 | 19.1 | 24.2 |
| Men | 2060 | 14.9 | 15.6 | 16.4 | 19.1 | 22.0 |
| **Total** | 2060 | 13.5 | 14.5 | 15.5 | 19.1 | 23.1 |
|  |  | **Type 2 diabetes cases** |  |  |  |  |
| Women | 2022 | 3,846,090 | 3,846,090 | 3,846,090 | 3,846,090 | 3,846,090 |
| Men | 2022 | 4,513,330 | 4,513,330 | 4,513,330 | 4,513,330 | 4,513,330 |
| **Total** | 2022 | 8,359,420 | 8,359,420 | 8,359,420 | 8,359,420 | 8,359,420 |
| Women | 2040 | 4,985,730 | 5,236,910 | 5,615,410 | 6,895,530 | 8,144,650 |
| Men | 2040 | 5,919,590 | 6,074,290 | 6,293,210 | 7,084,770 | 7,787,350 |
| **Total** | 2040 | 10,905,320 | 11,311,200 | 11,908,620 | 13,980,300 | 15,932,000 |
| Women | 2060 | 5,128,610 | 5,614,310 | 6,178,420 | 8,027,930 | 10,162,060 |
| Men | 2060 | 6,086,280 | 6,375,840 | 6,692,680 | 7,809,130 | 9,002,980 |
| **Total** | 2060 | 11,214,890 | 11,990,150 | 12,871,100 | 15,837,060 | 19,165,040 |

In addition to these general trends, a closer look at the prevalence and prevalence ratios (PRs) (Table 2) highlights the extent of the projected increase in the burden of type 2 diabetes between 2022 and 2060 when comparing the highest and lowest GISD groups.

**Table 2:** Prevalence in Germany in 2022, 2040 and 2060 by sex and highest and lowest quintile of the German Index of Socioeconomic Deprivation (GISD) and prevalence ratio between the highest and lowest GISD quintiles by sex, shown for all five scenarios. Decreasing and increasing scenarios show declining or rising type 2 diabetes incidence until 2027 (5-year de- or increase) and 2032 (10-year de- or increase), respectively, while the constant scenario assumes stable incidence from 2022 to 2060. Scenarios with increasing type 2 diabetes incidence extrapolate the non-linear trends observed between 2014 and 2022, capturing the recent increase, whereas decreasing scenarios extrapolate the long-term linear decrease in incidence.

| **GISD quintile** | **Sex** | **Calendar year** | **10-year decrease** | **5-year decrease** | **Constant incidence** | **5-year increase** | **10-year increase** |
|---|---|---|---|---|---|---|---|
|  |  |  | **Prevalence (%)** |  |  |  |  |
| **Lowest** | Women | 2022 | 7.9 | 7.9 | 7.9 | 7.9 | 7.9 |
|  |  | 2040 | 10.2 | 10.6 | 11.4 | 13.3 | 15.1 |
|  |  | 2060 | 10.8 | 11.7 | 12.9 | 15.8 | 19.1 |
|  | Men | 2022 | 9.8 | 9.8 | 9.8 | 9.8 | 9.8 |
|  |  | 2040 | 12.8 | 13.0 | 13.4 | 14.3 | 15.0 |
|  |  | 2060 | 13.4 | 14.0 | 14.6 | 15.9 | 17.2 |
| **Highest** | Women | 2022 | 10.4 | 10.4 | 10.4 | 10.4 | 10.4 |
|  |  | 2040 | 13.1 | 13.7 | 14.7 | 18.8 | 22.8 |
|  |  | 2060 | 13.6 | 14.9 | 16.5 | 22.3 | 29.1 |
|  | Men | 2022 | 12.2 | 12.2 | 12.2 | 12.2 | 12.2 |
|  |  | 2040 | 15.7 | 16.2 | 16.8 | 19.7 | 22.4 |
|  |  | 2060 | 16.3 | 17.2 | 18.1 | 22.2 | 26.7 |
|  |  |  | **Prevalence ratio, highest vs. lowest GISD group** |  |  |  |  |
|  | Women | 2022 | 1.31 | 1.31 | 1.31 | 1.31 | 1.31 |
|  |  | 2040 | 1.28 | 1.29 | 1.28 | 1.41 | 1.50 |
|  |  | 2060 | 1.25 | 1.27 | 1.27 | 1.41 | 1.52 |
|  | Men | 2022 | 1.24 | 1.24 | 1.24 | 1.24 | 1.24 |
|  |  | 2040 | 1.22 | 1.25 | 1.25 | 1.37 | 1.49 |
|  |  | 2060 | 1.21 | 1.23 | 1.24 | 1.39 | 1.55 |

The PRs reveal a growing relative disparity between the highest and lowest GISD quintile over the projection period. In 2022, women in the highest GISD group had a 31% higher prevalence compared to those in the lowest group. By 2060, this difference is projected to increase to 41% assuming a 5-year increase in incidence. A similar pattern was observed among men, with a ratio between the highest and lowest GISD quintiles increasing from 24% in 2022 to 39% in 2060.

Sensitivity analyses showed that, under the rather extreme scenarios considered, migration could have a substantial impact on projected type 2 diabetes prevalence and the future number of persons with the disease (Supplement 6–9). However, the projected socioeconomic inequalities remained largely unchanged (Supplement 9). As the actual prevalence among immigrants and emigrants is unknown, the extent and direction of any potential impact on our projections remain uncertain.

## DISCUSSION

Based on an illness-death-model using the most recently available representative data, we projected future age- and sex-specific type 2 diabetes prevalence and the number of persons with type 2 diabetes, as well as socioeconomic inequalities measured by the GISD, in Germany between 2022 and 2060. Across all considered projection scenarios, prevalence was projected to increase from 8.4 million in 2022 to between 11.2 and 19.2 million in 2060. Inequalities in prevalence tended to increase in scenarios with increasing type 2 diabetes incidence and were rather stable in scenarios with decreasing incidence, with increasing variation between GISD quintiles over time, particularly in scenarios with rising incidence.

### Comparison with previous studies

Our findings are in line with previous studies reporting increasing prevalence and number of persons with type 2 diabetes. Baumert et al.^17^ considered a total of 14 scenarios, with scenarios 1–5 corresponding to those described in Tönnies et al.^4^ Similar to our results, they projected higher prevalence of diagnosed type 2 diabetes by 2050, driven by declining excess mortality and, in particular, increasing incidence. Tönnies et al.^2^ projected future type 2 diabetes prevalence in Germany up to 2040. In each of the five applied scenarios, they assumed increasing type 2 diabetes incidence over different durations. Across all projection scenarios, an increase in the number of persons with type 2 diabetes was observed, with the magnitude of the increase depending on the duration of the assumed rise in incidence.

Both studies applied the same illness-death-model as used in our analysis. However, neither of them accounted for SEP. Despite this difference, their total numbers are comparable to ours, with previous projections estimating 10.9–14.2 million cases by 2040 and approximately 11.0 million cases by 2050. Beyond the national context, similar estimates from the Global Burden of Disease Study 2021^18^ have been published for other European countries. In small countries such as Malta, case numbers rose from approximately 11,300 cases in 1990 to 52,600 cases in 2021, respectively, while in larger countries such as France or Italy, cases increased by several million.^18^ This trend reflects a consistent rise in type 2 diabetes prevalence across the continent, highlighting the growing public health burden of the disease over the past three decades.

### Implications for future research and interventions for Germany

Our findings highlight the crucial role of incidence in determining future type 2 diabetes prevalence. Targeted prevention strategies could therefore substantially reduce future prevalence, particularly in socially deprived regions, underscoring the need for measures that address socioeconomic disparities. Such measures could include promoting healthier lifestyles, for example by adding more green spaces in deprived areas to encourage physical activity^19^, or by adjusting taxes on different foods^20^ to promote healthier dietary habits.^19^ The projected increase in type 2 diabetes prevalence in Germany implies a substantial increase in future healthcare needs. As the number of persons living with type 2 diabetes grows, so will the demand for medical care, diabetes management, and long-term monitoring.^21^ This trend is expected to place a significant financial burden on the healthcare system, given the high costs associated not only with diabetes itself but also with its numerous comorbidities, such as cardiovascular diseases or neuropathy.^22, 23^ Consequently, maintaining and improving the quality of care will be essential to prevent complications and ensure equitable access to effective treatment, particularly in light of persistent and increasing health inequalities.

Furthermore, demographic change will likely exacerbate these challenges: the German population is ageing, and older adults have a higher risk of developing type 2 diabetes and other chronic conditions. At the same time, the healthcare workforce is ageing as well, with many physicians approaching retirement age, which may further strain healthcare capacity.^24^ Overall, the combination of increasing Type 2 diabetes prevalence, population ageing, and limited healthcare resources underscores the urgent need for preventive measures and strategic healthcare planning to ensure sustainable diabetes care in the coming decades.

### Strength and limitations

A key strength of this study lies in the use of incidence trends derived from statutory health insurance data, which cover approximately 85% of the German population.^11^ The remaining 15% are either privately insured or, to a lesser extent, uninsured individuals. Private health insurance is generally associated with better health outcomes, higher SEP, and improved access to healthcare compared to statutory insurance.^25, 26^ Our findings therefore reflect the majority of the German population; however, as privately insured individuals are not included, projections for the total population may be slightly overestimated. Furthermore, the model captures the dynamic interplay between prevalence, incidence, and mortality, all of which vary by calendar year and age. In addition, the model can be extended to include socioeconomic differences as measured by the GISD. This is the first study, which includes the SEP in projections of future type 2 diabetes prevalence in Germany, allowing us to explicitly quantify socioeconomic inequalities across GISD quintiles and identify the particularly high future burden in highly deprived regions. Despite these strengths, the following limitations should be noted. Although the incidence trend used covers almost a decade, it is still too short to reliably estimate long-term developments. More robust assumptions about future trends would require data over a longer period. To address this limitation, we modelled different scenarios for future incidence trends. Determining which scenario is most likely, however, remains speculative.

The increase in type 2 diabetes incidence observed from 2019 onwards, particularly between 2020 and 2022, may be partly related to the COVID-19 pandemic. Although healthcare utilization declined during lockdown periods, lifestyle changes such as reduced physical activity, weight gain, and altered dietary habits may have contributed to a genuine rise in new type 2 diabetes cases.^27^ In addition, postponed routine check-ups and delayed diagnoses could have led to an apparent increase once healthcare access normalized. Therefore, the trend should continue to be monitored. Moreover, all projections exclusively refer to diagnosed type 2 diabetes, without considering the proportion of undiagnosed cases.

In all scenarios we used the temporal trend in MRR from Denmark^16^, but it would be more informative if German data on MRR were available. Moreover, the MRR is not stratified by GISD. However, this is expected to have a negligible effect, given that mortality exerts a smaller influence on future prevalence than incidence.^4^ Our projections do not account for changes in overall economic development. GISD quintiles and type 2 diabetes incidence were based on the 2019 distribution and assumed constant over the observation period. Consequently, potential socioeconomic changes are not reflected. Nevertheless, the GISD enables analyses of health inequalities in Germany when individual-level SEP data are unavailable. However, county-level GISD may not fully capture individual SEP.

## CONCLUSION

Overall, our findings indicate a substantial increase in type 2 diabetes prevalence in Germany between 2022 and 2060. The future burden will depend not only on the continuation of current incidence trends but also on sex and on SEP. Socioeconomic inequalities in prevalence are expected to widen if current trends persist, with highly deprived regions bearing a disproportionate future burden. The magnitude of this increase largely depends on how long upward trend in type 2 diabetes incidence continues. Therefore, careful monitoring of incidence trends in the years following the pandemic is essential. To prevent a further rise in socially patterned health inequalities, prevention and health promotion efforts should specifically target populations with low SEP.

## Funding

Funded by the Deutsche Forschungsgemeinschaft (DFG, German Research Foundation) – project number 516146656.

## Conflicts of Interest

The authors declare no conflicts of interest.

## Acknowledgements

We thank the Central Research Institute for Ambulatory Health Care in Germany (Zi) for sharing data used in this publication and for their support.

AI was used to shorten and to optimize text passages.

## Data Availability

The GISD data and a documentation of the data are public available under the following link: https://doi.org/10.5281/zenodo.6840304.

## Ethics approval

This study was conducted in accordance with the principles of the Declaration of Helsinki. The Ethics Committee of the Medical Faculty of Heinrich Heine University Düsseldorf has approved this study (study number: 2024-2722).

Only data that is routinely documented for billing purposes in the context of outpatient care, that has already been published, or that is made publicly available by the federal and state statistical offices will be used.

No individual data is available for this research project, meaning that no individuals can be identified. Consequently, no consent can be obtained from individuals.

## SUPPLEMENTAL MATERIAL

**Supplement 1:**
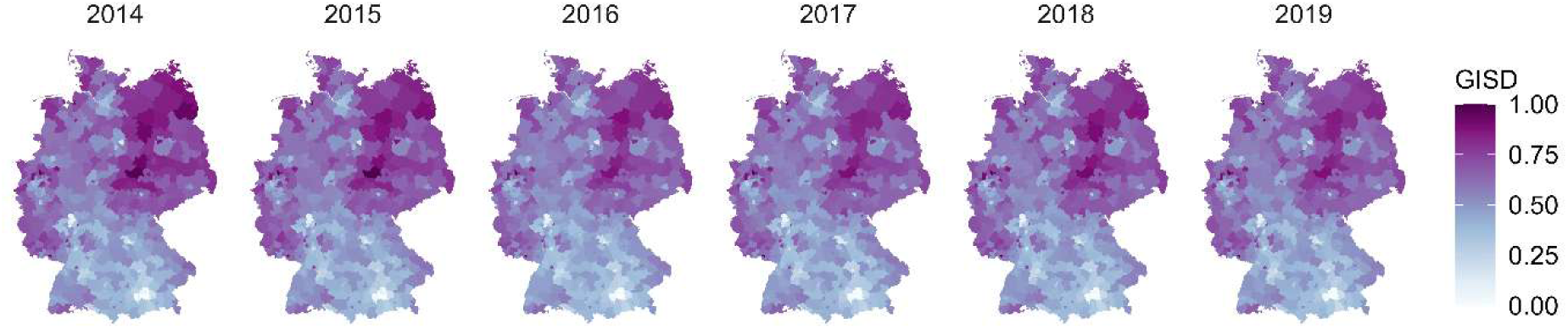
Distribution of the German Index of Socioeconomic Deprivation (GISD) between 2014 and 2019.

**Supplement 2:** Values for annual change in the type 2 diabetes incidence by sex and quintiles of the GISD used in the five scenarios for the projection of the future type 2 diabetes prevalence.

| **Scenario** | **GISD quintile** | **Annual change in the type 2 diabetes incidence (%)** |  |
|---|---|---|---|
|  |  | Men | Women |
| **Decrease in incidence** | 1 | -1.1 | -2.3 |
|  | 2 | -1.2 | -2.4 |
|  | 3 | -1.3 | -2.4 |
|  | 4 | -1.4 | -2.5 |
|  | 5 | -1.5 | -2.6 |
| **Constant incidence** | 1-5 | 0 | 0 |
| **Increase in incidence** | 1 | 2.3 | 5.3 |
|  | 2 | 3.8 | 6.7 |
|  | 3 | 4.4 | 7.2 |
|  | 4 | 4.9 | 7.7 |
|  | 5 | 6.2 | 8.8 |

**Supplement 3:**
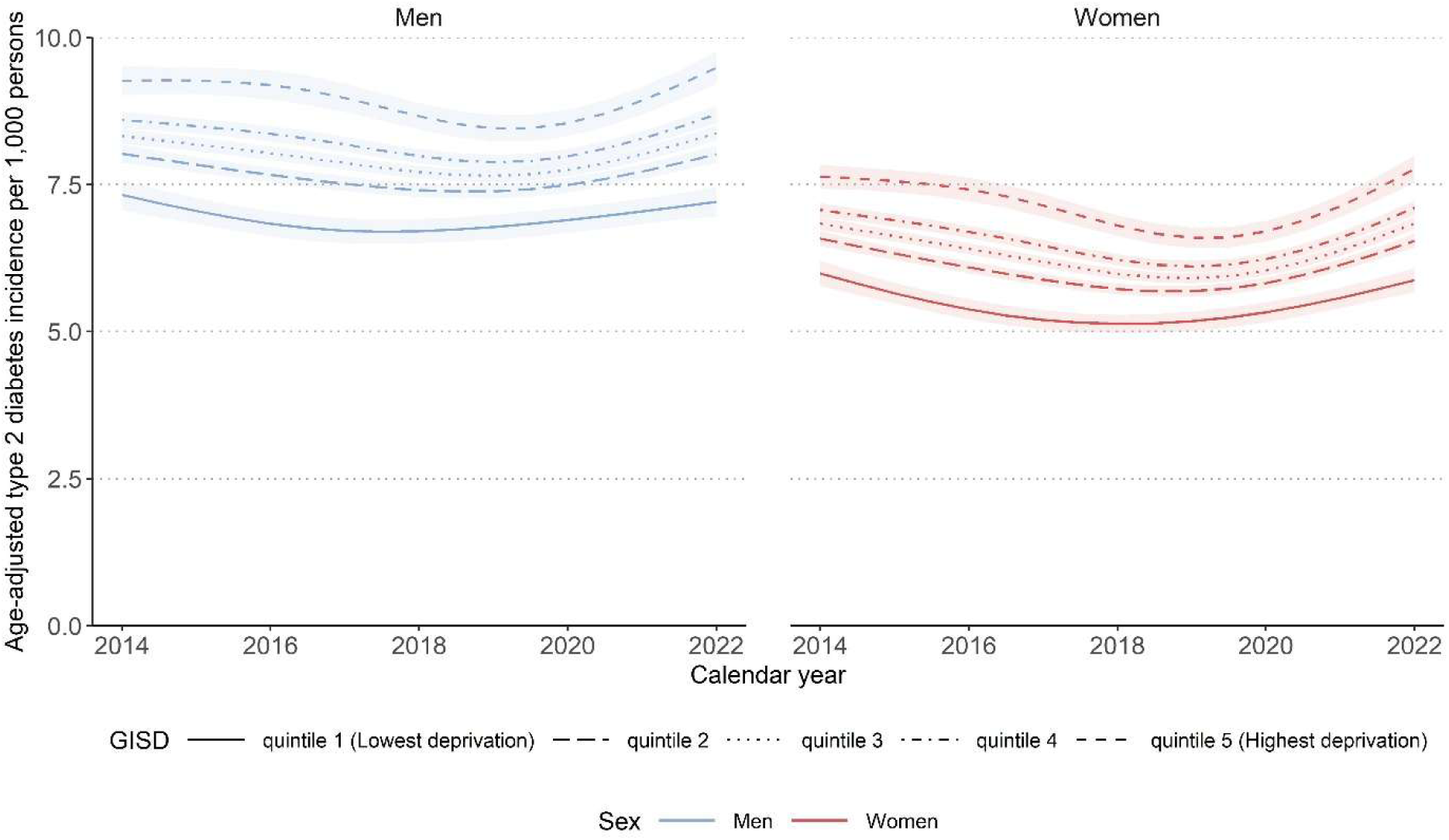
Age-adjusted incidence of type 2 diabetes (2014–2022) by quintiles of the German Index of Socioeconomic Deprivation (GISD). Results are based on a mixed negative binomial regression model. The model included GISD (linear), sex and calendar year (natural cubic spline with 3 degrees of freedom) as predictors, with the logarithm of expected cases as offset and counties as random intercepts. The outcome was the number of observed type 2 diabetes cases.

**Supplement 4:**
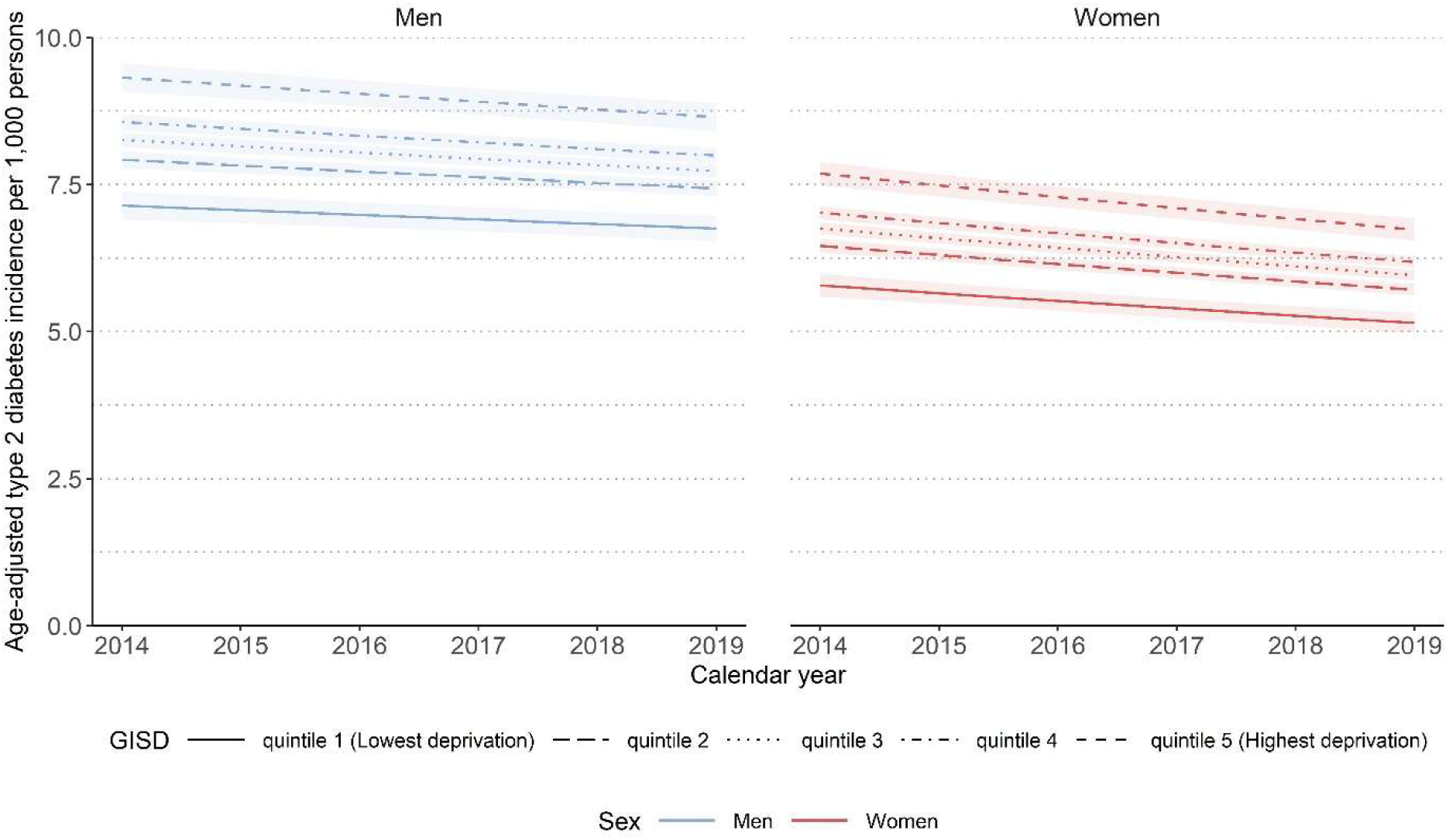
Age-adjusted incidence of type 2 diabetes (2014–2019) by quintiles of the German Index of Socioeconomic Deprivation (GISD). Results are based on a mixed negative binomial regression model. The model included GISD (linear), sex and calendar year (linear) as predictors, with the logarithm of expected cases as offset and counties as random intercepts. The outcome was the number of observed type 2 diabetes cases.

**Supplement 5:**
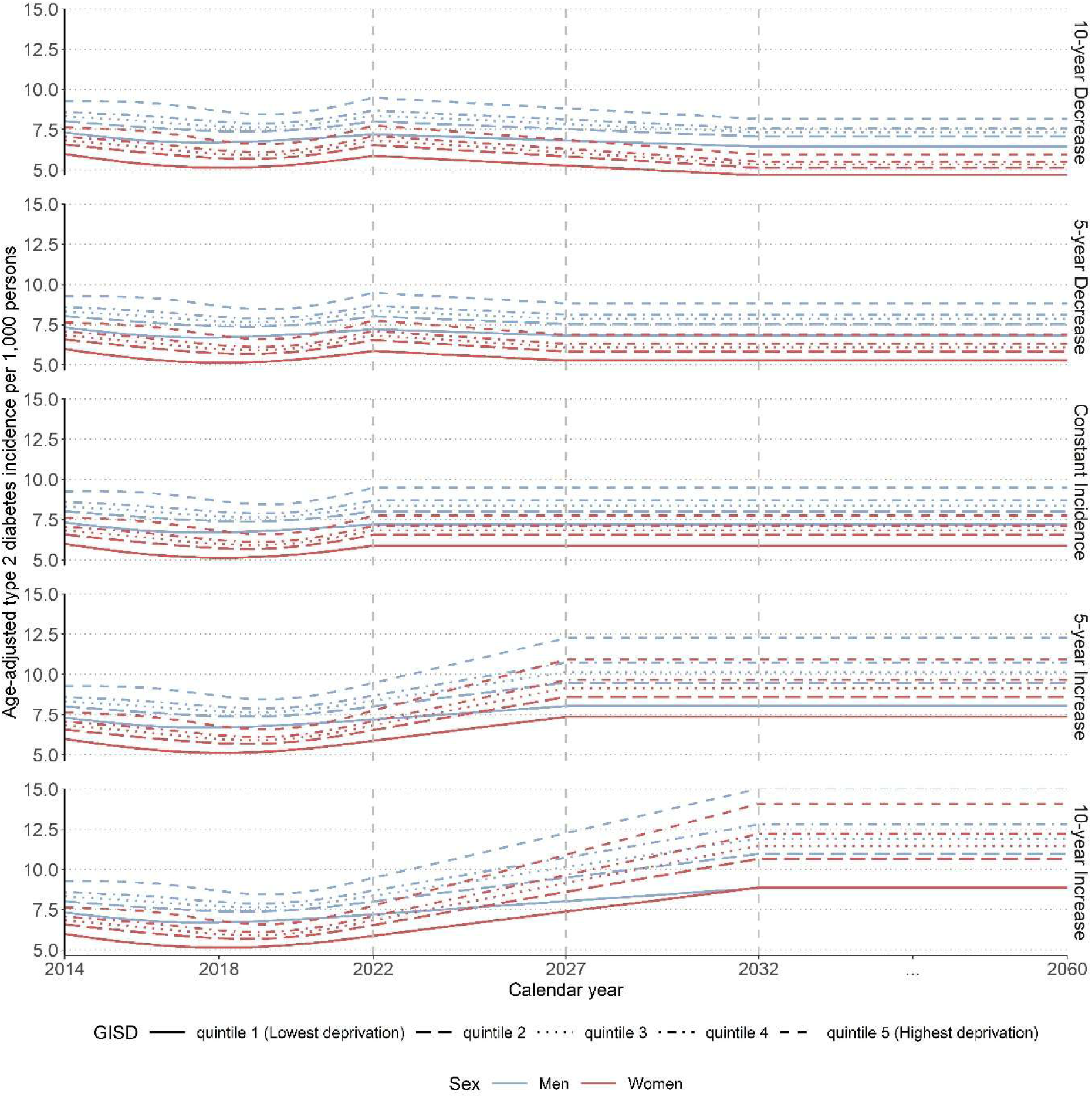
Visualization of all projection scenarios of future type 2 diabetes incidences in Germany between 2022 and 2060 among men (blue) and women (red) by quintiles of the German Index of Socioeconomic Deprivation (GISD). Decreasing and increasing scenarios show declining or rising type 2 diabetes incidence until 2027 (5-year in- or decrease) and 2032 (10-year in- or decrease), respectively, while the constant scenario assumes stable incidence from 2022 to 2060. Scenarios with increasing type 2 diabetes incidence extrapolate the non-linear trends observed between 2014 and 2022, capturing the recent increase, whereas decreasing scenarios extrapolate the long-term linear decrease in incidence.

**Supplement 6:**
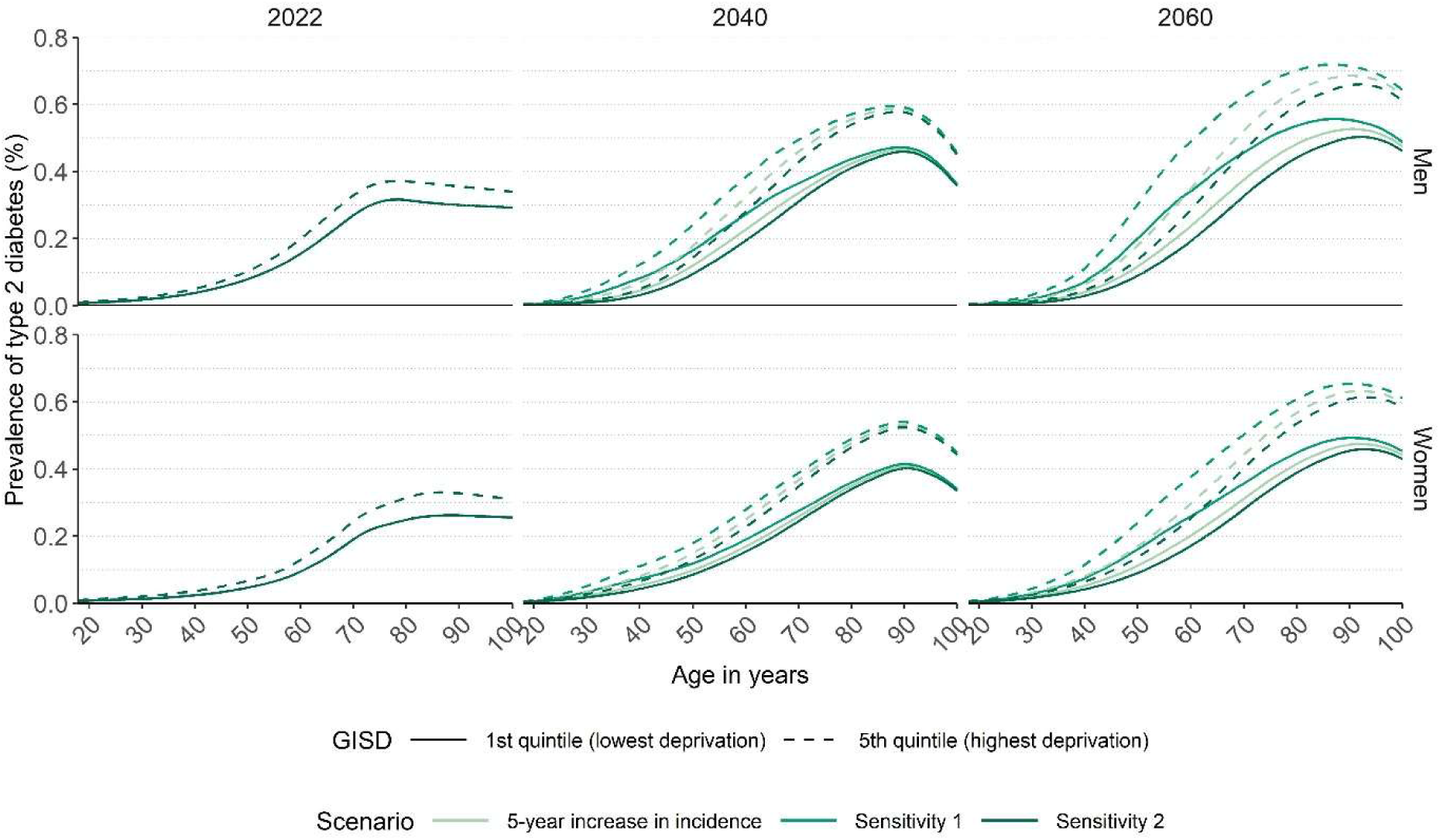
Projected age-specific prevalence of type 2 diabetes in Germany in 2022, 2040 and 2060 among men and women by the first and fifth quintile of the German Index of Socioeconomic Deprivation (GISD), shown for the scenario assuming a 5-year increase in type 2 diabetes incidence (see main text). The sensitivity analyses refer to different assumptions about the prevalence of migrants. In the first sensitivity analysis (sensitivity 1), we assume a threefold higher prevalence among persons immigrating to Germany and a prevalence one-third that of the resident population among persons emigrating from Germany. In the second sensitivity analysis (sensitivity 2), these assumptions are reversed, with a prevalence one-third that of the resident population among persons immigrating to Germany and a threefold higher prevalence among persons emigrating from Germany.

**Supplement 7:**
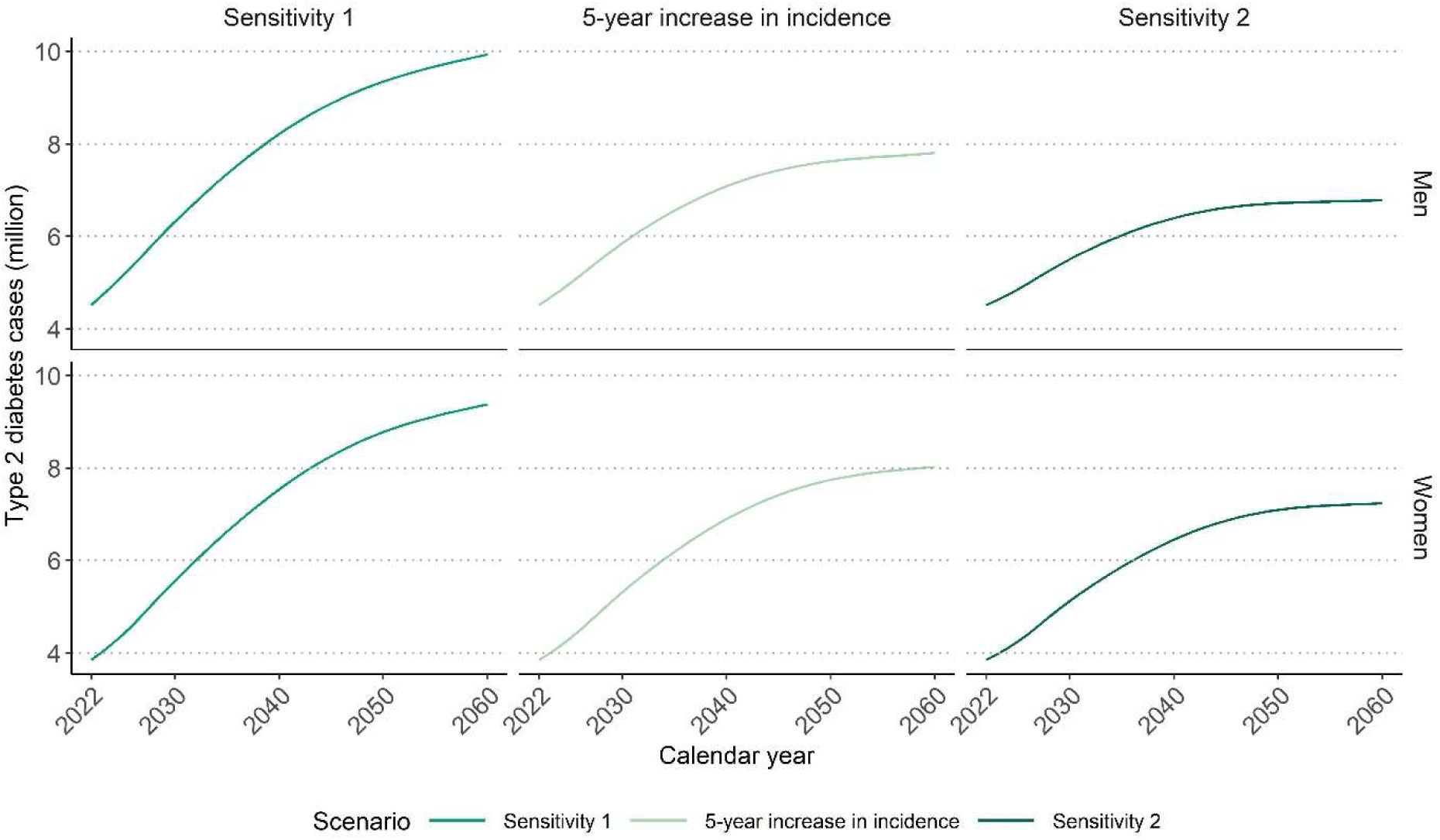
Projected number of type 2 diabetes cases in Germany between 2022 and 2060 summed across all quintiles of the German Index of Socioeconomic Deprivation (GISD), by sex shown for the scenario assuming a 5-year increase in type 2 diabetes incidence (see main text). The sensitivity analyses refer to different assumptions about the prevalence of migrants. In the first sensitivity analysis (sensitivity 1), we assume a threefold higher prevalence among persons immigrating to Germany and a prevalence one-third that of the resident population among persons emigrating from Germany. In the second sensitivity analysis (sensitivity 2), these assumptions are reversed, with a prevalence one-third that of the resident population among persons immigrating to Germany and a threefold higher prevalence among persons emigrating from Germany.

**Supplement 8:** Prevalence and total number of type 2 diabetes cases in Germany in 2022, 2040 and 2060 by sex. Projected age-specific prevalence of type 2 diabetes in Germany in 2022, 2040 and 2060 among men and women by quintiles of the German Index of Socioeconomic Deprivation (GISD), shown for the scenario assuming a 5-year increase in type 2 diabetes incidence (see main text). The sensitivity analyses refer to different assumptions about the prevalence of migrants. In the first sensitivity analysis (sensitivity 1), we assume a threefold higher prevalence among persons immigrating to Germany and a prevalence one-third that of the resident population among persons emigrating from Germany. In the second sensitivity analysis (sensitivity 2), these assumptions are reversed, with a prevalence one-third that of the resident population among persons immigrating to Germany and a threefold higher prevalence among persons emigrating from Germany.

| **Sex** | **Calendar year** | **Sensitivity 1** | **5-year increase** | **Sensitivity 2** |
|---|---|---|---|---|
|  |  | **Prevalence (%)** |  |  |
| Women | 2022 | 9.0 | 9.0 | 9.0 |
| Men | 2022 | 10.9 | 10.9 | 10.9 |
| **Total** | 2022 | 9.9 | 9.9 | 9.9 |
| Women | 2040 | 17.5 | 16.0 | 14.9 |
| Men | 2040 | 19.7 | 17.0 | 15.3 |
| **Total** | 2040 | 18.6 | 16.5 | 15.1 |
| Women | 2060 | 22.3 | 19.1 | 17.2 |
| Men | 2060 | 24.3 | 19.1 | 16.6 |
| **Total** | 2060 | 23.3 | 19.1 | 16.9 |
|  |  | **Type 2 diabetes cases** |  |  |
| Women | 2022 | 3,846,090 | 3,846,090 | 3,846,090 |
| Men | 2022 | 4,513,330 | 4,513,330 | 4,513,330 |
| **Total** | 2022 | 8,359,420 | 8,359,420 | 8,359,420 |
| Women | 2040 | 7,537,420 | 6,895,530 | 6,450,330 |
| Men | 2040 | 8,214,440 | 7,084,770 | 6,396,490 |
| **Total** | 2040 | 15,751,860 | 13,980,300 | 12,846,820 |
| Women | 2060 | 9,377,410 | 8,027,930 | 7,237,190 |
| Men | 2060 | 9,940,130 | 7,809,130 | 6,789,580 |
| **Total** | 2060 | 19,317,540 | 15,837,060 | 14,026,770 |

**Supplement 9:**
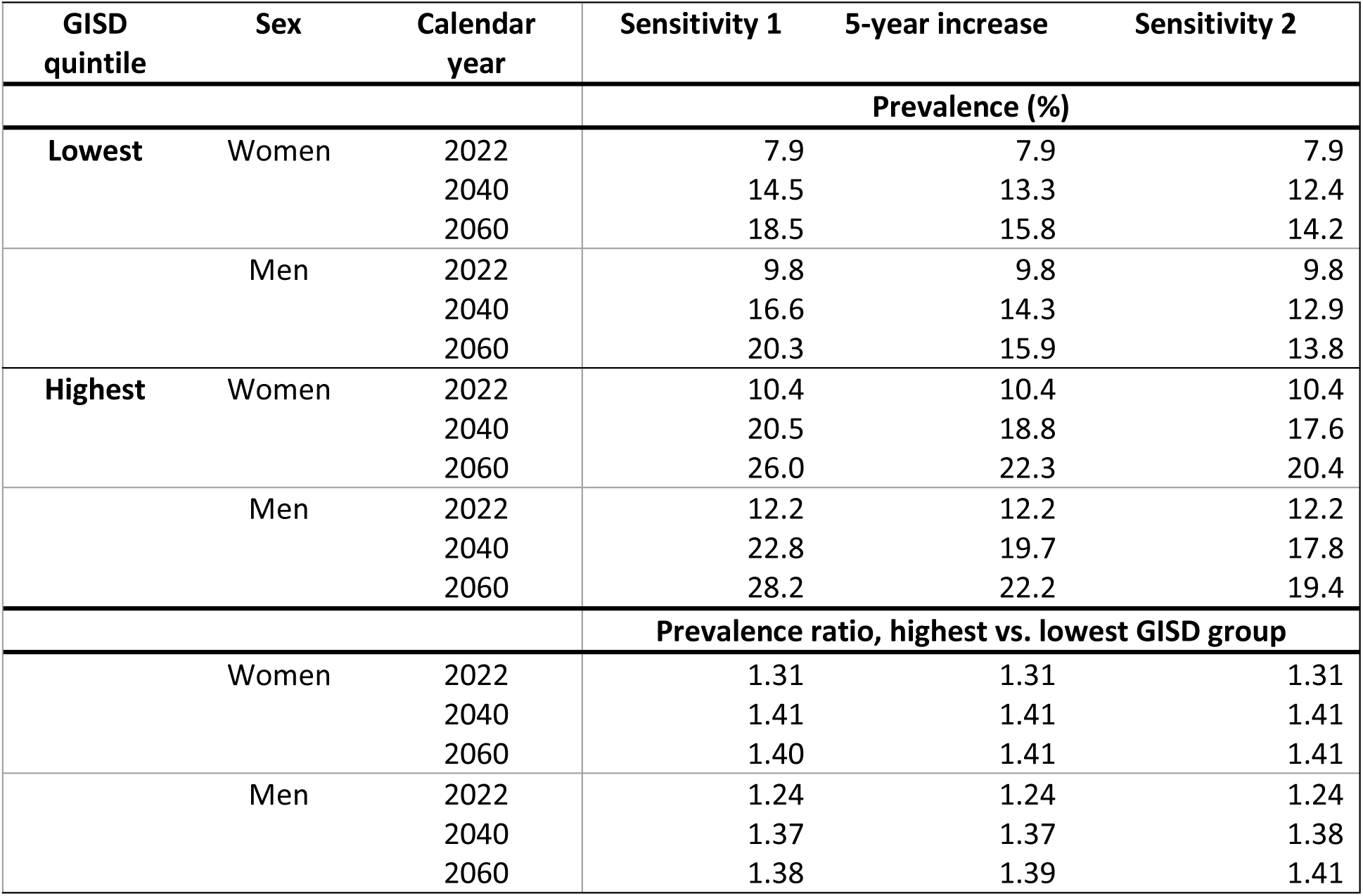
Prevalence in Germany in 2022, 2040 and 2060 by sex and highest and lowest quintile of the German Index of Socioeconomic Deprivation (GISD) and prevalence ratio between the highest and lowest GISD quintiles by sex, shown for the scenario assuming a 5-year increase in type 2 diabetes incidence (see main text). The sensitivity analyses refer to different assumptions about the prevalence of migrants. In the first sensitivity analysis (sensitivity 1), we assume a threefold higher prevalence among persons immigrating to Germany and a prevalence one-third that of the resident population among persons emigrating from Germany. In the second sensitivity analysis (sensitivity 2), these assumptions are reversed, with a prevalence one-third that of the resident population among persons immigrating to Germany and a threefold higher prevalence among persons emigrating from Germany.

| GISD quintile | Sex | Calendar year | Sensitivity 1 | 5-year increase | Sensitivity 2 |
|---|---|---|---|---|---|
|  |  |  | Prevalence (%) |  |  |
| **Lowest** | Women | 2022 | 7.9 | 7.9 | 7.9 |
|  |  | 2040 | 14.5 | 13.3 | 12.4 |
|  |  | 2060 | 18.5 | 15.8 | 14.2 |
|  | Men | 2022 | 9.8 | 9.8 | 9.8 |
|  |  | 2040 | 16.6 | 14.3 | 12.9 |
|  |  | 2060 | 20.3 | 15.9 | 13.8 |
| **Highest** | Women | 2022 | 10.4 | 10.4 | 10.4 |
|  |  | 2040 | 20.5 | 18.8 | 17.6 |
|  |  | 2060 | 26.0 | 22.3 | 20.4 |
|  | Men | 2022 | 12.2 | 12.2 | 12.2 |
|  |  | 2040 | 22.8 | 19.7 | 17.8 |
|  |  | 2060 | 28.2 | 22.2 | 19.4 |
|  |  |  | Prevalence ratio, highest vs. lowest GISD group |  |  |
|  | Women | 2022 | 1.31 | 1.31 | 1.31 |
|  |  | 2040 | 1.41 | 1.41 | 1.41 |
|  |  | 2060 | 1.40 | 1.41 | 1.41 |
|  | Men | 2022 | 1.24 | 1.24 | 1.24 |
|  |  | 2040 | 1.37 | 1.37 | 1.38 |
|  |  | 2060 | 1.38 | 1.39 | 1.41 |

## Notes

### Competing Interest Statement

The authors have declared no competing interest.

### Author Declarations

This study was conducted in accordance with the principles of the Declaration of Helsinki. The Ethics Committee of the Medical Faculty of Heinrich Heine University Duesseldorf has approved this study (study number: 2024-2722). Only data that is routinely documented for billing purposes in the context of outpatient care, that has already been published, or that is made publicly available by the federal and state statistical offices will be used. No individual data is available for this research project, meaning that no individuals can be identified. Consequently, no consent can be obtained from individuals.

